# Asymptomatic SARS-CoV-2 infection and the demography of COVID-19

**DOI:** 10.1101/2021.09.03.21262757

**Authors:** Zengmiao Wang, Peiyi Wu, Jingyuan Wang, José Lourenço, Bingying Li, Benjamin Rader, Marko Laine, Hui Miao, Ligui Wang, Hongbin Song, Nita Bharti, John S. Brownstein, Ottar N. Bjornstad, Christopher Dye, Huaiyu Tian

**Affiliations:** State Key Laboratory of Remote Sensing Science, Center for Global Change and Public Health, College of Global Change and Earth System Science, Beijing Normal University, Beijing, China; School of Computer Science and Engineering, Beihang University, China; Peng Cheng Laboratory, Shenzhen, China; Department of Zoology, University of Oxford, Oxford, UK; Computational Epidemiology Lab, Boston Children’s Hospital, Boston MA, United States; Department of Epidemiology, Boston University School of Public Health, Boston MA, USA; Finnish Meteorological Institute, Meteorological Research Unit, Helsinki, Finland; Department of Statistics, College of Art and Science, Ohio State University, Columbus, Ohio, USA; Center of Disease Control and Prevention, PLA, Beijing, China; Center for Infectious Disease Dynamics, Department of Biology, Pennsylvania State University, University Park, Pennsylvania, USA; Harvard Medical School, Harvard University, Boston, MA, USA; Department of Entomology, College of Agricultural Sciences, Pennsylvania State University, University Park, Pennsylvania, USA

## Abstract

Asymptomatic individuals carrying SARS-CoV-2 can transmit the virus and contribute to outbreaks of COVID-19, but it is not yet clear how the proportion of asymptomatic infections varies by age and geographic location. Here we use detailed surveillance data gathered during COVID-19 resurgences in six cities of China at the beginning of 2021 to investigate this question. Data were collected by multiple rounds of city-wide PCR test with detailed contact tracing, where each patient was monitored for symptoms through the whole course of infection. We find that the proportion of asymptomatic infections declines with age (coefficient =-0.006, *P*<0.01), falling from 56% in age group 0–9 years to 12% in age group >60 years. Using an age-stratified compartment model, we show that this age-dependent asymptomatic pattern together with the age distribution of overall cases can explain most of the geographic differences in reported asymptomatic proportions. Combined with demography and contact matrices from other countries worldwide, we estimate that a maximum of 22%–55% of SARS-CoV-2 infections would come from asymptomatic cases in an uncontrolled epidemic based on asymptomatic proportions in China. Our analysis suggests that flare-ups of COVID-19 are likely if only adults are vaccinated and that surveillance and possibly control measures among children will be still needed in the future to contain epidemic resurgence.

## Main text

An important clinical feature for SARS-CoV-2, distinguished from SARS ^1^, is that it spreads not only through symptomatic individuals but also infected individuals who are pre-symptomatic or asymptomatic ^2–4^. With the pandemic ongoing, the importance of asymptomatic cases is increasingly recognized ^5,6^ as these potentially undetected cases may trigger flare-ups circulating in the general community^7,8^. Since such a public health threat was recognized ^7,9,10^, numerous studies have been performed to decipher the underlying mechanism ^11,12,21,13–20^ and quantify the major drivers ^22,23^ of asymptomatic infection.

With the accumulating evidence of the role of asymptomatic infections, one surprising global feature is crystalizing: the proportion of asymptomatic infections show vast differences across the world ^24–26^. Studies have reported the proportion of asymptomatic cases varying from 1.21% to 91.88% (including estimates from serological screening studies, Figure 1A and B). Once age-stratified, the fraction of asymptomatic cases also show substantial variation (Figure 1C) ^27–29^ in age. This indicates that local demography may play a role in the observed geographical and age-related heterogeneity of asymptomatic proportions. The main obstacles to better understand this heterogeneity include measurement of asymptomatic status ^30,31^, testing bias ^10,23^ and sampling bias (for example, testing rates biased to hospitalized individuals ^32,33^). Because passive surveillance is often limited to symptomatic cases, a substantial proportion of asymptomatic cases are expected to be underreported ^34,35^. Asymptomatic transmission poses a great challenge to local governments and the health systems ^34,36–41^, in terms of control policy, being necessary to identify the factors driving such heterogeneity in asymptomatic proportions to more efficiently implement control that includes such subclass of the infected. For example, the local government may need to prepare for the mass-testing to accelerate the case identification and cut the transmission if the asymptomatic proportion is high due to the related factors.

**Figure 1.**
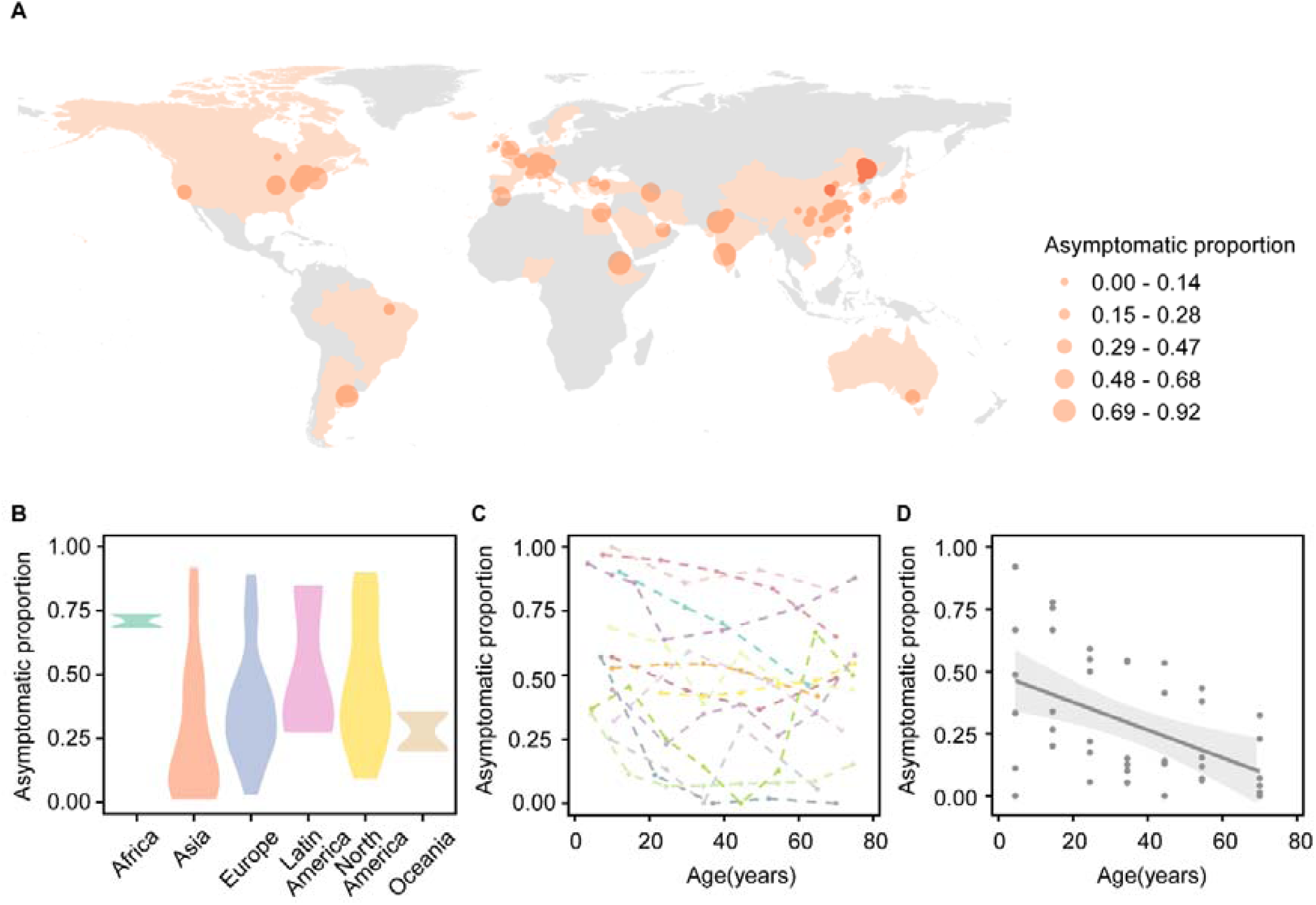
The asymptomatic SARS-CoV-2 cases in the world. (A) The dots on the maps indicate the location of the reported asymptomatic proportions. The dot size indicates the asymptomatic proportion. The red dot represents the data in our study. (B) The violin plot for the reported asymptomatic proportion in the continents. (C) The asymptomatic proportion reported by age in previous studies. (D) The grey points represent the data from Chinese cities. A linear regression was built (Asymptomatic proportion = Intercept+Coeffecient×Age). The grey line represents the linear regression using pooled asymptomatic proportion under each age group across cities with 95% CI in light grey shadow (Coefficient=-0.006, *P*-value<0.01). To get a reliable estimate, only the asymptomatic proportions derived from more than 50 samples are plotted. Note that the testing methods and definition of asymptomatic cases in these studies were different from ours. The asymptomatic proportion from other parts of the world may not be comparable to our data directly.

To address this question, exhaustive, active surveillance of COVID-19 epidemics is needed. Detailed surveillance data collected during COVID-19 resurgences in six cities of China at the beginning of 2021 provide a valuable opportunity to study this issue. Multiple rounds of population-level PCR tests in each city accelerated the speed of SARS-CoV-2 infection identification (the cases have short time interval between the infection and detection) and improved the detection rate among the currently infected irrespective of clinical outcome. All identified SARS-CoV-2 infection went into isolation. Detailed contact tracing and surveillance made it possible to define the clinical endpoint (asymptomatic or symptomatic) by monitoring the health status for patients and their contacts through the course of infection. Here we calculated the crude asymptomatic proportion as the number of SARS-CoV-2 carriers without any symptoms divided by the total number of SARS-CoV-2 carriers. Patients who showed no symptoms at testing and developed symptoms later were counted as symptomatic cases. Using this unique dataset, we explored the potential factors that may contribute to observed variations in the proportion of asymptomatic infections, and reports thereof. Our analysis has useful implications for understanding epidemic trajectories and can help guide future disease control policies that include the asymptomatic and potentially hidden subgroup of infections.

### The proportion of asymptomatic infections from multiple rounds of population-level PCR tests with patient follow-up

We collected data on SARS-CoV-2 infections from six cities in China (Shijiazhuang and Xingtai from Hebei province, Changchun and Tonghua from Jilin province, Harbin and Suihua from Heilongjiang province). The population size for the six cities is approximately 9.8 million, 8.0 million, 8.6 million, 2.1 million, 9.5 million and 5.2 million, respectively. To detect all ongoing infections as early as possible, the cities launched population-level PCR tests with more than 48 million tests in total (the actual number of tests should be higher than this since not all the information about testing could be collected for this study.); for example, Shijiazhuang performed 3 rounds of testing and Xingtai conducted as many as 10 rounds (Supplementary Table 1).

In total, there were 2,744 PCR-confirmed SARS-CoV-2 infections with the highest number of 1,040 in Shijiazhuang and the lowest number of 80 in Xingtai (Supplementary Table 1). The age distribution of infections varied by location: the largest proportion of positive cases was in individuals aged >60 in Shijiazhuang, Changchun, Tonghua and Suihua, while the largest proportion was in age group of 20-29 for Xingtai and in age group of 40-49 for Harbin (Supplementary Figure 1).

The total or “crude” asymptomatic proportion (i.e. the asymptomatic SARS-CoV2 infections divided by all the SARS-CoV2 infections for each city, see Methods) varied considerably, from 8% in Tonghua to 18% in Shijiazhuang and 51% in Harbin (white bars with black borders in Figure 2B). The bias in this asymptomatic proportion calculation should be very small due to following reasons: (1) the populations were tested multiple times during the COVID-19 flare-ups; (2) the identified cases were isolated and under 14-days health monitoring to create a definite clinical outcome (asymptomatic or symptomatic) per individual. Across the six cities, the proportion of asymptomatic infections declined with age (Figure 1D, coefficient =-0.006 and *P*<0.01 from linear regression, effect of gender and random effect of city in terms of intercept are accounted for).

**Figure 2.**
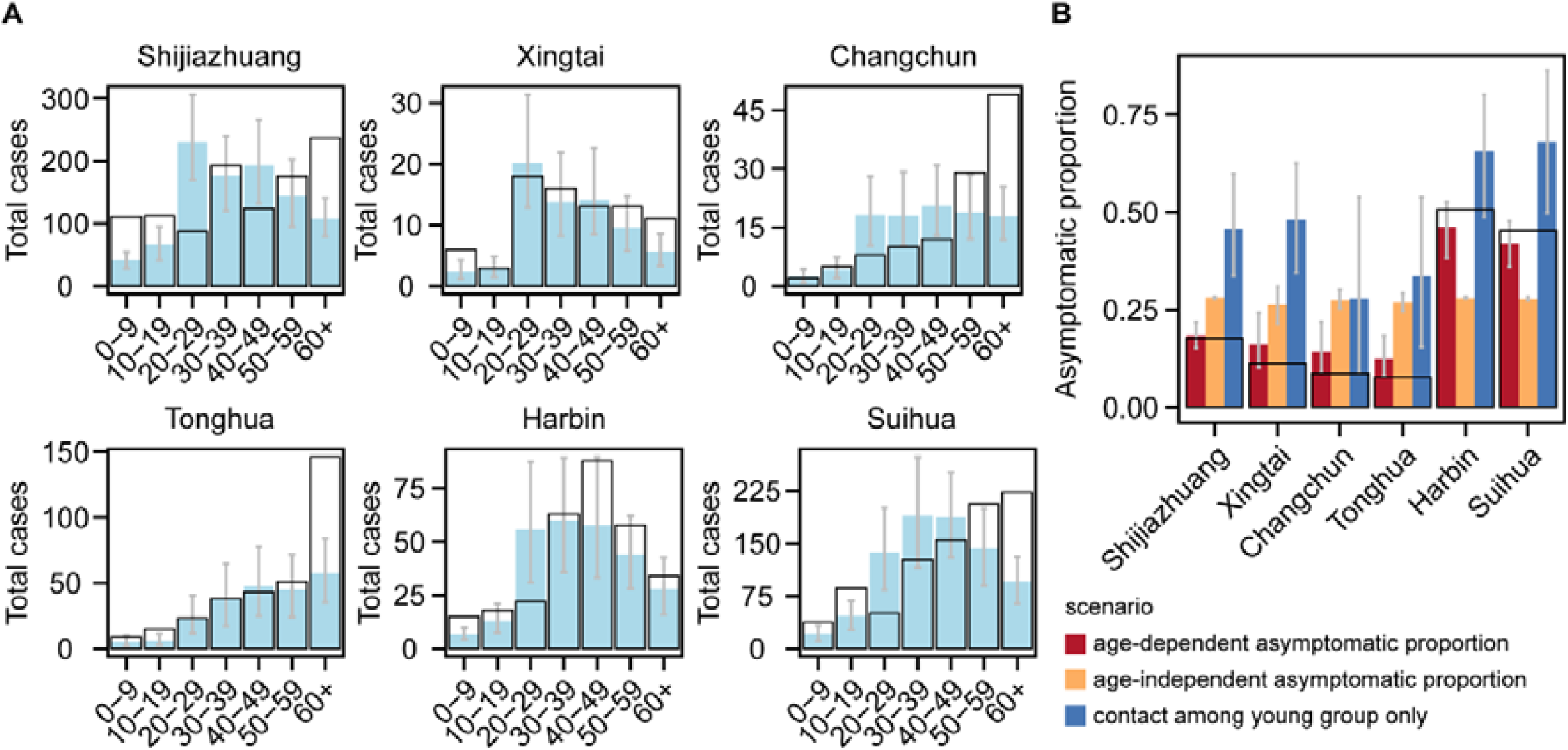
The estimated SARS-CoV-2 cases distribution in age groups in 6 cities and the total asymptomatic proportions under different scenarios. (A) Estimate of the developed model to the total number of SARS-CoV-2 cases in age groups for 6 cities. The total number of SARS-CoV-2 cases reported (black box) and the estimated total number of cases (light blue box) in each city and each age group. The grey line represents 95% CI. (B) The total asymptomatic proportions under different scenarios. Red: simulated, based on the observed age-dependent asymptomatic proportions. Yellow: simulated, with age-independent asymptomatic proportions. This proportion was calculated by the total asymptomatic cases divided by the total SARS-CoV-2 cases using the pooled data from 6 cities. Blue: simulated, with contacts among the 0-9 and 10-19 age group only. The grey bar represents the 95% CI for each simulation. White bars with black borders represent the observed total asymptomatic proportion in each city.

Some studies have reported that COVID-19 vaccines lower the risk of being symptomatic ^42–44^. However, vaccines are unlikely to contribute to heterogeneity of asymptomatic proportion in our study because the vaccine roll-out had not started at the time of data collection. It is also unlikely that exposure profiles induced by the first COVID-19 wave contributed to the heterogeneity of asymptomatic proportions in this data set, because the total number of SARS-CoV-2 infections during the first wave in 2020 among these 6 cities was small: ranging from 7 in Tonghua to 198 in Harbin. Additionally, a retrospective serological survey conducted in 6 provinces or municipalities (out of Hubei province) from April 10, 2020 to April 18, 2020 suggested that less than 0.1% of the population carried antibodies against SARS-CoV-2 ^45^.

### Age-dependent asymptomatic proportions and epidemic trajectories

We hypothesized that cities report different total asymptomatic proportions of SARS-CoV-2 infections because of an interplay between age-dependent asymptomatic proportion of cases, the age distribution of infections and the age distribution of the underlying susceptible population. To explore this hypothesis, we developed an age-stratified discrete stochastic compartment model that incorporated age-dependent asymptomatic proportions to reconstruct observed epidemic trajectories including both the asymptomatic and symptomatic cases (Supplementary Figure 2). The proportion of asymptomatic infections was assumed to decline with age, in accordance to our findings (Figure 1D) and other report ^46^. The daily numbers of asymptomatic and symptomatic cases (Supplementary Figure 3) were well predicted. Overall, the model predicted the case distribution in age (Figure 2A), the age-dependent proportion of asymptomatic cases (Supplementary Figure 4), and the reported asymptomatic proportion in all cases (red bar in Figure 2B) across all the cities (note that these were not involved in the loss function during fitting).

Based on the fit of the model, we next investigated the effect of the age-dependent asymptomatic proportions on epidemic trajectories. Our model suggested that if the proportion of asymptomatic cases had been the same across all age groups (i.e. the age-independent), the total proportion of asymptomatic cases in each city would have been equal (yellow bar in Figure 2B and Supplementary Figure 5). However, this contradicted the observations and illustrated that the asymptomatic proportion depends on age. The results demonstrated that age-dependent asymptomatic patterns and the age distribution of cases explain some but not all of the heterogeneity between geographic locations.

Vaccines are rolling out among adults. However, the future epidemic trajectories are unclear when all adults are vaccinated. To imitate an epidemic where adults are vaccinated and children are not vaccinated, we simulated a scenario in which sustained transmission occurred only among the young population (contacts assumed only to take place among age groups [0-9] and [10-19]). Such a scenario assumed that all adults (>19 years old) are vaccinated, and that vaccines are 100% effective against infection. The total asymptomatic proportion was predicted to be higher than observed (blue bar in Figure 2B and Supplementary Figure 6), because of the shifted age distribution of cases (Supplementary Figure 7A-7D). These results illustrate that even in scenarios where all adults are vaccinated and protected from infection, sustained symptomatic flare-ups of COVID-19 among the young are possible. Surveillance and control measures among younger age groups may be still needed to prevent this

Using the observed pattern from China paired with globally comprehensive covariates, we simulated SARS-CoV-2 epidemic trajectories for 150 countries across all continents (i.e. 150 nation-level datasets). Results showed that asymptomatic cases contribute to SARS-CoV-2 transmission anywhere from 6.3% to 22.1% (Figure 3A, 20.8%-55.1% under the assumption of largest asymptomatic proportion for each age group, Supplementary Figure 8A) and the total asymptomatic proportion may be from 13.9 to 47.9% (Figure 3B, 40.2%-80.5% under the assumption of largest asymptomatic proportion for each age group, Supplementary Figure 8B). The national median age determines the contribution of asymptomatic infections and the total asymptomatic proportion (Figure 3D and 3E, Supplementary Figure 8D and 8E), but these factors do not influence the time to the peak of the epidemic (Figure 3C and 3F, Supplementary Figure 8C and 8F). Age-dependent asymptomatic patterns and population demographics may indicate the risk of repeated waves at the global scale in an uncontrolled situation; and countries with younger populations may face a high risk of COVID-19 outbreaks due to undetected transmission. However, cultural and socioeconomic differences among countries are likely to greatly influence our base-line projections.

**Figure 3.**
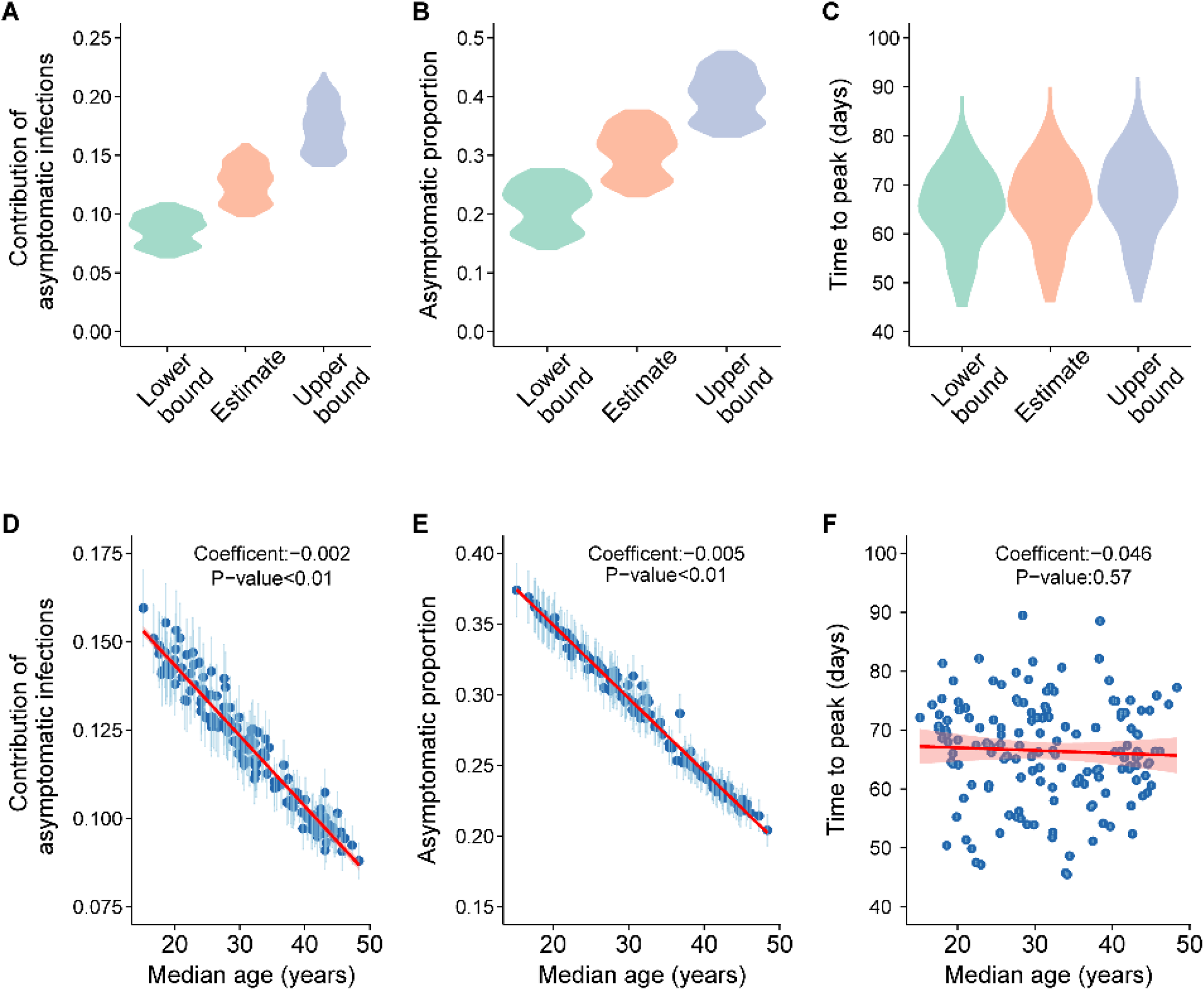
Application to other 150 countries in the world for an uncontrolled epidemic. (A) Contribution to SARS-CoV-2 infections by asymptomatic infections of age-dependent asymptomatic proportion for 150 countries. The contribution of asymptomatic infection is defined as the percentage of SARS-CoV-2 cases infected by the asymptomatic cases. A linear regression was built between age groups and asymptomatic proportion (Figure 1C). Using the fitted linear model, the estimates and the confidence interval of asymptomatic proportion under each age group were obtained. Estimate: the estimated asymptomatic proportions for 7 age groups (the grey line in Figure 1C). Upper bound and lower bound: 95% confidence interval of asymptomatic proportion for each age group (the grey shadow in Figure 1C). (B) Total asymptomatic proportion for 150 countries. (C) Time to peak for 150 countries. (D) The association between the contribution of asymptomatic infections and the median age across 150 countries. For each country, the transmission rate was scaled to keep *R*_0_ =2.4. We simulated 20 outbreaks in each country by drawing the age-dependent asymptomatic proportion randomly from the confidence interval provided in Figure 1C. The blue dots represent the average with 95% CI in light blue across 20 simulations. A linear regression was fitted (red line) between median age of a given country and (D) contribution of asymptomatic infection, (E) the total asymptomatic proportion, and (F) time to peak.

Our study provides insight into the observed variability in the proportion of asymptomatic SARS-CoV-2 infections. Together, local age-dependent asymptomatic proportions along with the age distribution of cases lead to differences in observed asymptomatic proportions. Based on previous ^10,29,47–49^ studies and the data from our study, the proportion of asymptomatic cases as a non-increasing function of age is supported. However, future studies are needed to understand how asymptomatic proportions will change in the face of accumulated natural exposure, emerging variants, and immunity induced by imperfect vaccines. Our results do clarify how COVID-19 flare-ups may happen from largely asymptomatic, susceptible and unvaccinated young age groups. Surveillance and control measures among young age groups in the near future may still be needed to avoid resurgence. In summary, our study may provide some insights for control of future resurgences. The age-dependent asymptomatic proportions may affect the epidemic trajectory and age-group specific control measures may be needed to combat this SARS-CoV-2 virus.

## Methods

### The definition for asymptomatic and symptomatic SARS-CoV-2 cases Suspected COVID-19 case

Individuals who have any one of the epidemiological histories, and meet any two of the clinical manifestations; if there is no clear epidemiological history, meet any 2 of the clinical manifestations, and the SARS-CoV-2-specific IgM antibody is positive; or meets 3 of the clinical manifestations.

#### Epidemiological history

1) There was travel history or residence history of the community with SARS-CoV-2 cases up to 14 days before the onset of the disease; 2) A history of contact with a case or asymptomatic infection up to 14 days before the onset of the disease; 3) Contact with fever and/or respiratory symptoms from the community with reported COVID-19 cases in the 14 days before the onset of illness; 4) Cluster disease: 2 or more cases of fever and/or respiratory symptoms occurred in a small area (such as homes, offices, school classes, etc.) within 14 days.

#### Clinical manifestations

1) Fever or respiratory symptoms; 2) Radiographic evidence of COVID-19; 3) The total number of white blood cells is normal or decreased, and the lymphocyte count is normal or decreased during early onset.

### Symptomatic SARS-CoV-2 cases

Suspected cases have one of the following etiological or serological evidences: (1) real-time fluorescent RT-PCR detection of SARS-CoV-2 nucleic acid positive; (2) viral genome sequencing, which is highly homologous to known SARS-CoV-2; (3) SARS-CoV-2-specific IgM antibody and IgG antibody are both positive; (4) SARS-CoV-2-specific IgG antibody turns from negative to positive or the IgG antibody titer in the recovery phase is 4 times or more higher than that in the acute phase.

### Asymptomatic SARS-CoV-2 cases

Respiratory tract and other specimens tested positive using PCR or viral genome sequencing for the pathogen of SARS-CoV-2 without relevant clinical manifestations, such as fever, dry cough, sore throat, and other self-perceived symptoms or clinically recognizable signs, and CT imaging without features of COVID-19. After 14 days of medical observation in quarantine, there were no self-perceived symptoms or clinically identifiable signs.

### The SARS-CoV-2 case data

For each city, the detailed surveillance data was collected from local Centers for Disease Control and Prevention, including the gender, age, and onset date. For each patient, the clinical endpoint (asymptomatic or symptomatic) was also collected over a multi-week period.

### The global analysis

The country-specific contact matrices were obtained from previous study^50^. The population demographics for 2020 were obtained from World Population Prospects 2019 and 150 countries were included in this analysis.

### The age-dependent asymptomatic proportion and the total asymptomatic proportion

Based on the observed data, the asymptomatic proportion is dependent on age. Let *p*_*1*_, *…, p*_*7*_ represent the asymptomatic proportion in age group 0-9, 10-19, 20-29, 30-39, 40-49, 50-59, 60+, respectively. So, the total asymptomatic proportion (*p*) is calculated as following:

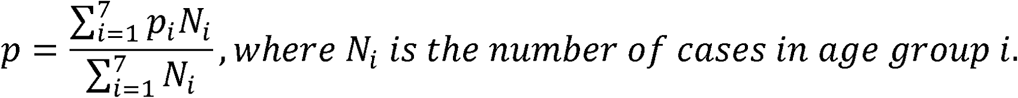

The total asymptomatic proportion is a combination of the age-dependent asymptomatic proportion and the SARS-CoV-2 case distribution in each age groups.

### The mobility data

In China, human movements were anonymously collected at the city level with mobile phone data, through location-based services (LBS) employed by the popular Baidu applications. We used relative volume of inflows movement for each city from the migration flows database (http://qianxi.baidu.com/). The study period for Shijangzhuang, Xingtai, Changchun, Tonghua, Harbin and Suihua was Dec 23, 2020-Feb 12, 2021, Jan 1, 2021-Jan 24, 2021, Jan 10, 2021-Feb 4, 2021, Jan 15, 2021-Feb 9, 2021, Jan 6, 2021-Feb 8, 2021 and Jan 9, 2021-Feb 5, 2021, respectively.

### The population-level PCR tests and the contact tracing

The rounds of testing and the total number of tests performed in each city were collected from news sources. For the procedure of PCR tests and contact tracing, please see the Supplementary Information.

## Data Availability

The data supporting the findings of this study are available within the article.

## Author contributions

H.T. and Z.W. designed the study. J.W., B.L., H.M., L.W., and H.S. collected the statistical data. Z.W. and P.W. conducted the analyses. J.L., M.L., B.R., N.B., J.S., and O.N.B. edited the manuscript. Z.W., H.T., and C.D. wrote the manuscript. All authors read and approved the manuscript.

## Data and materials availability

The data supporting the findings of this study are available within the article.

## Ethics approval and consent to participate

The epidemiological and clinical data collection was exempt from Institutional Review Boards, because it was part of public health investigation for COVID-19, issued by the National Health Commission of the People’s Republic of China (http://www.nhc.gov.cn/jkj/s3577/202003/4856d5b0458141fa9f376853224d41d7.shtml). This study was approved by the Institutional Review Boards from the Chinese Center for Disease Control and Prevention.

