## Supplementary Information for "Asymptomatic SARS-CoV-2 infection and the demography of COVID-19"

This PDF file includes:

Supplementary text

References for SI reference citations

**Materials and Methods**

**Summary of contact and contact tracing**

***Shijiazhuang:*** As of January 8^th^, 7,864 close contacts and 3,581 secondary close contacts were screened, and 3,857 people were quarantined in a centralized manner (<https://baijiahao.baidu.com/s?id=1688314189476367095&wfr=spider&for=pc>).

***Xingtai:*** On January 4^th^, 2021, 1,092 close contacts of confirmed cases and 929 close contacts of close contacts were screened (<http://m.news.cctv.com/2021/01/06/ARTIRR5moe33P2faCXFCXks4210106.shtml>).

***Changchun***: As of 24:00 on February 1, a total of 237,391 people was traced, of which 2,375 were close contacts, 5,514 were close contacts of close contacts, and 229,502 were from other high-risk groups. A total of 5,241 people were quarantined in a centralized manner, and 105,612 people were isolated at home (<http://www.scio.gov.cn/xwfbh/gssxwfbh/xwfbh/jilin/Document/1698273/1698273.htm>).

***Tonghua***: As of 24:00 on February 1, a total of 3,076 close contacts and 4,214 close contacts of close contacts have been investigated (<http://www.scio.gov.cn/xwfbh/gssxwfbh/xwfbh/jilin/Document/1698273/1698273.htm>).

***Harbin:*** From January 9^th^ to February 4^th^ 24:00, a total of 4,947 close contacts and 21,231 close contacts of close contacts were traced ([https://mbd.baidu.com/newspage/data/landingsuper?third=baijiahao&baijiahao_id=1690867370715866615&c_source=kunlun&p_tk=46681L5JdDdRUjwmpxxxZdrmRxWBOQQG%2F9uP7Dg6SlvHuVXCj0QmSU35%2F1c1nZUszU4JMyrw37C4yYOHM4XRKs0anw06fEhVvYHchp4gEKsEVo3r3suJKENYghkB1bTRdx4f](https://baijiahao.baidu.com/s?id=1688314189476367095&wfr=spider&for=pc)).

***Suihua:*** The contacts data was not found.

**Definition for contacts**

The definition of close contacts: individuals who are in short physical distance with suspected cases and confirmed cases starting 2 days before the onset of symptoms, or 2 days before sampling of asymptomatic infections, but who have not taken effective protection.

***Determination of close contacts***:

1. Family members living together in the same room;

2. Direct caregivers or provide diagnosis, treatment and nursing services;

3. Medical staff who carry out diagnostic and treatment activities that may generate aerosols in the same space

4. Individuals who have a short physical distance in offices, workshops, teams, elevators, canteens, classrooms, etc.

5. Those who share meals, entertain together, and provide catering and entertainment services in a confined environment.

6. Medical staff, family members, or other close contacts visiting the case.

7. People who take the same transportation and have close contact (within 1 meter), including caregivers and accompanying persons (family, colleagues, friends, etc.) on the transportation.

8. Exposure to persons who may be contaminated by cases or asymptomatic infections.

9. Individuals who were assessed by the on-site investigators based on the local circumstances.

The definition of close contact of close contact: people who have close contact with the first contact (the person who has close contact with cases), such as living together, working in the same closed environment, gathering meals, and entertainment, but have not taken effective protection. The close contact with the first contact should be taken place during the interval from the first contact in a short physical distance with the SARS-CoV-2 infections to the isolation of the first contact.

General contact definition: people who have had contact with suspected cases, confirmed cases, and asymptomatic infected persons who have been in contact with the same means of transportation, living together, studying, working, and diagnosis and treatment such as airplanes, trains, ships, etc., but do not meet the principle of determining close contacts.

**Contact tracing**

A field epidemiology investigation was performed by the local CDC when a SARS-CoV-2 infection was identified. Activity patterns of the case starting 14 days before symptom onset and until confirmation or isolation were collected. The case was also asked to provide a list of she/he visited and their contacts. With this information, the contact tracing was launched, including the interviews with contacts, checking the travel records and the digital health records. If an individual meets the definition of close contact, she/he will be quarantined.

**The PCR tests**

***Nasopharyngeal swab***: The sampler gently holds the person's head with one hand, the swab in another, insert the swab via nostril to enter, slowly get deep along the bottom of the lower nasal canal. Because the nasal canal is curved, do not force too hard to avoid traumatic bleeding. When the tip of the swab reaches the posterior wall of the nasopharyngeal cavity, rotate gently once (pause for a moment in case of reflex cough), then slowly remove the swab and dip the swab tip into a tube containing 2-3ml virus preservation solution (or isotonic saline solution, tissue culture solution or phosphate buffer), discard the tail and tighten the cap.

***Pharynx swab***: the sampled person first gargles with normal saline, the sampler immerses the swabs in sterile saline (virus preservation solution is not allowed to avoid antibiotic allergies), holds the head of the sampled person up slightly, with one’s mouth wide open, making a sound "ah" to expose the lateral pharyngeal tonsils, insert the swabs, stick across the tongue roots, and wipe both sides of the pharyngeal tonsils with pressure at least 3 times, then wipe on the upper and lower walls of the pharynxes for at least 3 times, and dip the swabs in a tube containing 2-3ml storage solution (or isotonic saline solution, tissue culture solution or phosphate buffer solution), discard the tail and tighten the cap. The pharyngeal swabs can also be placed in the same tube together with the nasopharyngeal swab.

***Novel coronavirus nucleic acid assay (real-time fluorescence-based RT-PCR assay)***: Primers and probes target the ORF1ab and N gene regions of the novel coronavirus. For nucleic acid extraction and real-time fluorescence-based RT-PCR reaction system and reaction conditions, kit instructions of the manufacturers were followed.

***Judgment of results:***

Negative: no Ct value or Ct value≥ 40.

Positive: Ct value <37.

Gray zone: Repeated experiments are recommended should Ct value range between 37 and 40. If the Ct value reads <40 and the amplification curve has obvious peaks, the sample should be considered being tested positive, otherwise it should be considered as negative.

Note: If a commercial kit is used, the instructions provided by the manufacturer shall prevail.
